# Detection of Infectious Corneal Perforation Using Anterior Segment Optical Coherence Tomography

**DOI:** 10.64898/2026.01.28.26345085

**Authors:** Folahan Ibukun, Kamini Reddy, Subeesh Kuyyadiyil, Elesh Jain, Gautam Parmar, Nakul S. Shekhawat

## Abstract

**Purpose:** To evaluate the diagnostic performance of anterior segment optical coherence tomography (ASOCT) compared to slit lamp examination for identification of corneal perforation in microbial keratitis, and to assess ASOCT grading reproducibility.

**Methods:** We conducted a diagnostic concordance study of 150 eyes with microbial keratitis at a tertiary eye hospital in India. Two masked graders independently evaluated ASOCT scans for perforation, with disagreements resolved by consensus. We calculated Cohen’s kappa for inter-grader concordance and intra-grader repeatability. Sensitivity and specificity of slit lamp examination were calculated using ASOCT as the reference standard. Logistic regression identified factors associated with disagreement between modalities.

**Results:** Inter-grader agreement for ASOCT was near-perfect (κ=0.98; 95% CI, 0.92-1.00). ASOCT identified perforation in 24 eyes (16.0%) compared to 12 eyes (8.0%) by slit lamp examination. Using ASOCT as reference, slit lamp examination demonstrated 33.3% sensitivity (95% CI, 14.9-52.2%) and 96.8% specificity (95% CI, 93.4-99.2%). Odds of disagreement were significantly higher for eyes with stromal thinning (OR=8.19; 95% CI, 2.27-29.54), mid-stromal involvement (OR=4.44; 95% CI, 1.08-18.30), and infection within 2mm of the limbus (OR=8.81; 95% CI, 1.77-43.80).

**Conclusions:** ASOCT enables highly reproducible perforation grading and detects substantially more perforations than slit lamp examination, particularly in severe disease. These findings support ASOCT as an objective tool for clinical assessment and outcome ascertainment in microbial keratitis.

## INTRODUCTION

Microbial keratitis is the leading cause of corneal blindness worldwide and ranks among the top causes of visual impairment globally.^1,2^ The disease burden is particularly severe in low- and middle-income countries, where agricultural occupations, limited access to eye care, and delayed presentation contribute to poor visual outcomes.^3,4^ Corneal perforation represents one of the most devastating complications of microbial keratitis as it may lead to profound corneal damage, prolapse of intraocular contents, endophthalmitis, and ultimately loss of the eye.^5,6^ Management options for perforation depend on its size, location, and anatomic structure, and range from observation with medical therapy alone – for small perforations with iris plugging that is sufficient to maintain a formed anterior chamber – to cyanoacrylate gluing,^7^ Tenon patch graft,^8^ multilayered amniotic membrane grafting,^9,10^ corneal tissue patch graft, mini-DSAEK,^11^ or tectonic penetrating keratoplasty.^12,13^ Accurate identification of the presence, location, extent, and anatomic structure of corneal perforation is therefore essential for corneal surgeons to determine the necessity, optimal method, and timing of perforation repair.

In clinical research studies evaluating outcomes of microbial keratitis, perforation is frequently defined as an outcome or adverse event indicating treatment failure. For example, the Mycotic Ulcer Treatment Trial demonstrated that topical natamycin was superior to topical voriconazole for treatment of filamentous fungal keratitis based not only on visual acuity outcomes but also on reduced occurrence of perforation or therapeutic keratoplasty.^14,15^ The reliability of perforation as an endpoint therefore has direct implications for the ability of clinical trials to detect treatment effects and generate evidence to guide clinical practice.

On slit lamp examination, corneal perforation is typically identified via direct visualization of the stromal defect, iris tissue plugging the perforation, or a shallow or collapsed anterior chamber. Seidel testing with fluorescein can confirm the diagnosis and identify micro-perforations that might be missed on routine examination. However, densely opaque corneas with stromal infiltrates or scarring can make it difficult to visualize perforations or gauge anterior chamber depth, and even Seidel testing can be negative when there is sufficient iris plugging of the perforation. The phenotypic spectrum of perforation ranges from subtle micro-perforations with minimal aqueous leak to frank iris prolapse through large corneal defects, introducing substantial subjectivity to clinical assessment. Detection of corneal perforation may be even more difficult when remotely reviewing slit lamp photographs, which could reduce the utility of anterior segment photography for clinical trial reading centers and telemedicine monitoring.

Anterior segment optical coherence tomography (ASOCT) offers an imaging-based approach to evaluate corneal pathology with high-resolution cross-sectional visualization.^16–19^ Newer ASOCT modalities with high wavelength penetration allow visualization of deeper tissue and can delineate stromal architecture, quantify corneal thickness, and identify structural abnormalities including descemetoceles and perforations that may not be readily apparent on slit lamp examination.^16,20–25^ ASOCT could enable more objective, standardized, and reproducible detection of perforation detection than subjective clinical examination, with potential applications both in clinical decision-making and as an outcome measure for clinical trials. However, the diagnostic utility of ASOCT for corneal perforation in microbial keratitis has not been formally evaluated.

The purpose of this study was to evaluate the diagnostic performance of ASOCT and slit lamp photography compared to in-person slit lamp examination for identification of corneal perforation in microbial keratitis, and to assess the intra-grader and inter-grader concordance of ASOCT-based perforation grading.

## METHODS

### Study Design and Population

We conducted a prospective diagnostic concordance study among consecutive patients with microbiologically confirmed microbial keratitis treated at SNC Chitrakoot, a tertiary eye care center in Madhya Pradesh, India, between May 2024 and January 2025. Eligible patients included those with bacterial, fungal, or polymicrobial keratitis confirmed by microbiologic culture or smear testing. We excluded eyes with ungradable image quality, unreadable ASOCT files, or missing slit lamp examination findings. All participants provided informed consent prior to study enrollment. This study adhered to the tenets of the Declaration of Helsinki and received approval from the SNC Chitrakoot Eye Hospital institutional ethics committee and the Johns Hopkins University School of Medicine institutional review board.

### In-Person Slit Lamp Examination

All patients underwent a comprehensive ophthalmic examination including best-corrected visual acuity measurement and slit lamp biomicroscopy. The examining ophthalmologist, masked to ASOCT and slit lamp photography findings, documented the presence or absence of frank corneal perforation on slit lamp examination along with detailed characterization of slit lamp examination findings including epithelial defect size, infiltrate or scar diameter and depth, and infiltrate or scar centrality. The presence and extent of stromal thinning was also recorded. ASOCT imaging and slit lamp photography were performed in a separate location from ophthalmologists’ slit lamp examination to ensure that clinical findings did not influence image acquisition or interpretation.

### Photography and ASOCT Imaging

Slit lamp photographs with diffuse illumination as well as cobalt blue lighting following fluorescein instillation were obtained. ASOCT imaging was performed using the Heidelberg Anterion platform (Heidelberg Engineering, Heidelberg, Germany) with the Metrics App, which obtains six radial cross-sectional scans that capture limbus-to-limbus anatomy of the cornea and anterior segment. This standardized imaging protocol provided comprehensive sampling of the corneal architecture across multiple evenly spaced meridians, enabling visualization of a majority of the anterior segment including any areas of stromal pathology, corneal thinning, iris abnormality, or anterior chamber shallowing. For each case, a four-panel composite image was created comprising the ASOCT scan, a diffuse light slit lamp photograph, a cobalt blue light slit lamp photograph, and the ASOCT angle indicator superimposed on a grayscale photograph of the cornea to assist with anatomic orientation (Figure 1). When slit lamp photographs were unavailable, smartphone photographs of comparable quality were substituted.

**Figure 1.**
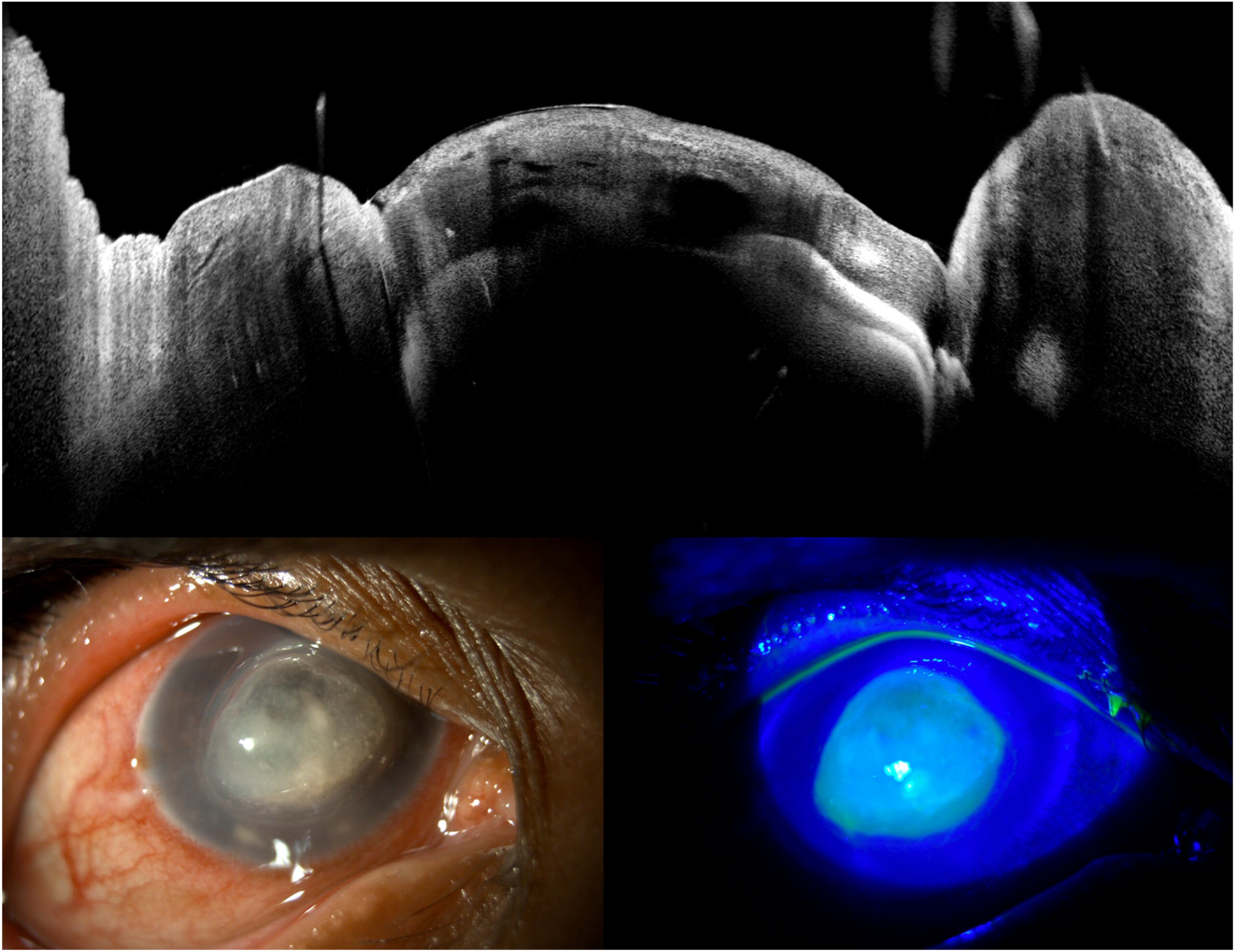
Example of corneal perforation on ASOCT versus slit lamp photography. Although slit lamp examination and masked graders evaluating slit lamp photographs did not identify perforation, both masked graders identified perforation upon evaluating the ASOCT scan.

### Image Grading

For each eye, two masked ophthalmologist graders (NS, FI) independently evaluated all six radial ASOCT images alongside slit lamp photographs and graded the eye for presence or absence of frank corneal perforation. Graders were masked to one another’s assessments as well as to the in-person clinical diagnosis. Frank corneal perforation was defined as a full-thickness defect of the stroma with Descemet membrane discontinuity, or a collapsed or nearly collapsed anterior chamber associated with evidence of severe corneal thinning and/or descemetocele. For this initial analysis focused on frank perforation, shallow anterior chamber depth due to leakage of fluid through an impending perforation or descemetocele without frank perforation did not qualify. Shallow anterior chambers associated with iridocorneal adhesions, synechiae, or other iridocorneal angle abnormalities were not classified as perforation unless there was overt evidence of iris tissue plugging an area of perforated cornea. Old perforations that had healed with re-epithelialization over the area of iris plugging were also not classified as frank or active perforation. Following independent grading, if the two graders disagreed on the presence or absence of corneal perforation for an eye, both graders viewed and discussed that eye’s images together until a consensus assessment was achieved. To assess intra-grader reproducibility, a randomly selected 30% subsample of the study population was designated for repeat grading by both graders at a separate session. When completing repeat grading, graders were masked to their own previous assessments as well as each other’s assessments.

In a separate image reading session, the two graders also independently and remotely evaluated slit lamp photographs alone (without paired ASOCT). Slit lamp photographs consisted of one diffuse illumination photograph and one blue light photograph taken 15 seconds after instillation of fluorescein dye. Slit lamp photograph grading was performed using the same masked grading and adjudication protocol as for ASOCT composite images. We assessed intra- and inter-grader agreement of remote slit lamp photography grading using a similar approach as for ASOCT composite images.

### Comparing Diagnostic Performance Across Modalities

We evaluated diagnostic performance across three pairwise comparisons: (1) ASOCT versus in-person slit lamp examination; (2) Slit lamp photography versus in-person slit lamp examination; and (3) ASOCT versus slit lamp photography. For comparisons involving ASOCT, consensus ASOCT grading served as the reference standard. For comparison of slit lamp photography to in-person slit lamp examination, the latter served as the reference standard.

### Statistical Analysis

Intra-grader reproducibility was quantified using Cohen’s kappa statistic and percent agreement for the repeated assessments within the 30% subsample. Inter-grader agreement between the two graders was similarly assessed using percent agreement and Cohen’s kappa for the binary classification of “present” versus “absent” perforation. We report the proportion of images requiring adjudication due to initial disagreement and the final distribution of consensus diagnoses.

To evaluate diagnostic validity, we calculated sensitivity, specificity, positive predictive value, negative predictive value, and overall percent agreement of the tested modality relative to the reference standard with 95% percentile-based confidence intervals derived using a bootstrap approach with 1,000 resampling iterations.

To identify clinical factors associated with agreement versus disagreement between ASOCT and in-person examination, we performed univariable and multivariable logistic regression analyses with robust variance. Covariates examined in multivariable modeling included visual acuity (logarithm of minimum angle of resolution [logMAR] <1.0 vs. ≥1.0), infection type (bacterial, fungal, or polymicrobial), infection size (stromal infiltrate diameter <2mm, 2-6mm, or ≥6mm), infiltrate depth, presence of stromal thinning and proximity to limbus (within 2mm). P values of <0.05 were considered statistically significant. All statistical analyses were performed using Stata SE version 18.5 (StataCorp, College Station, TX).

## RESULTS

### Study Population

Of 167 eyes from 162 patients initially screened, 150 eyes (N=96 right eye [64.0%]) from 150 patients met eligibility criteria and were included in the analysis (Table 1). Exclusions included 10 eyes with ungradable image quality, 4 eyes incorrectly marked as having active infection, 2 eyes with unreadable ASOCT files, and 1 eye with missing in-person slit lamp grading information. Among the 150 included eyes, 5 did not have ASOCT scans obtained at the baseline visit. For these eyes, scans from the next consecutive visit were used for composite image creation. Overall, 92 patients (61.3%) were male. The median age was 50.0 years (interquartile range [IQR]= 41.0–59.0). By sex, the median was 49.0 years for females (IQR= 43.0–54.0) and 51.0 years for males (IQR= 41.0–62.5). Mean logMAR visual acuity was 1.43 (standard deviation= 0.86). Fifty-one eyes (34.0%) had light perception only, and one eye (0.7%) had no light perception. The median time from symptom onset to hospital presentation was 15 days (IQR= 7–30 days). The study population comprised patients with fungal keratitis (n=80, 53.3%), bacterial keratitis (n=54, 36.0%), and polymicrobial keratitis (n=16, 10.7%). Additional clinical characteristics of the study population are presented in Table 1.

**Table 1.**
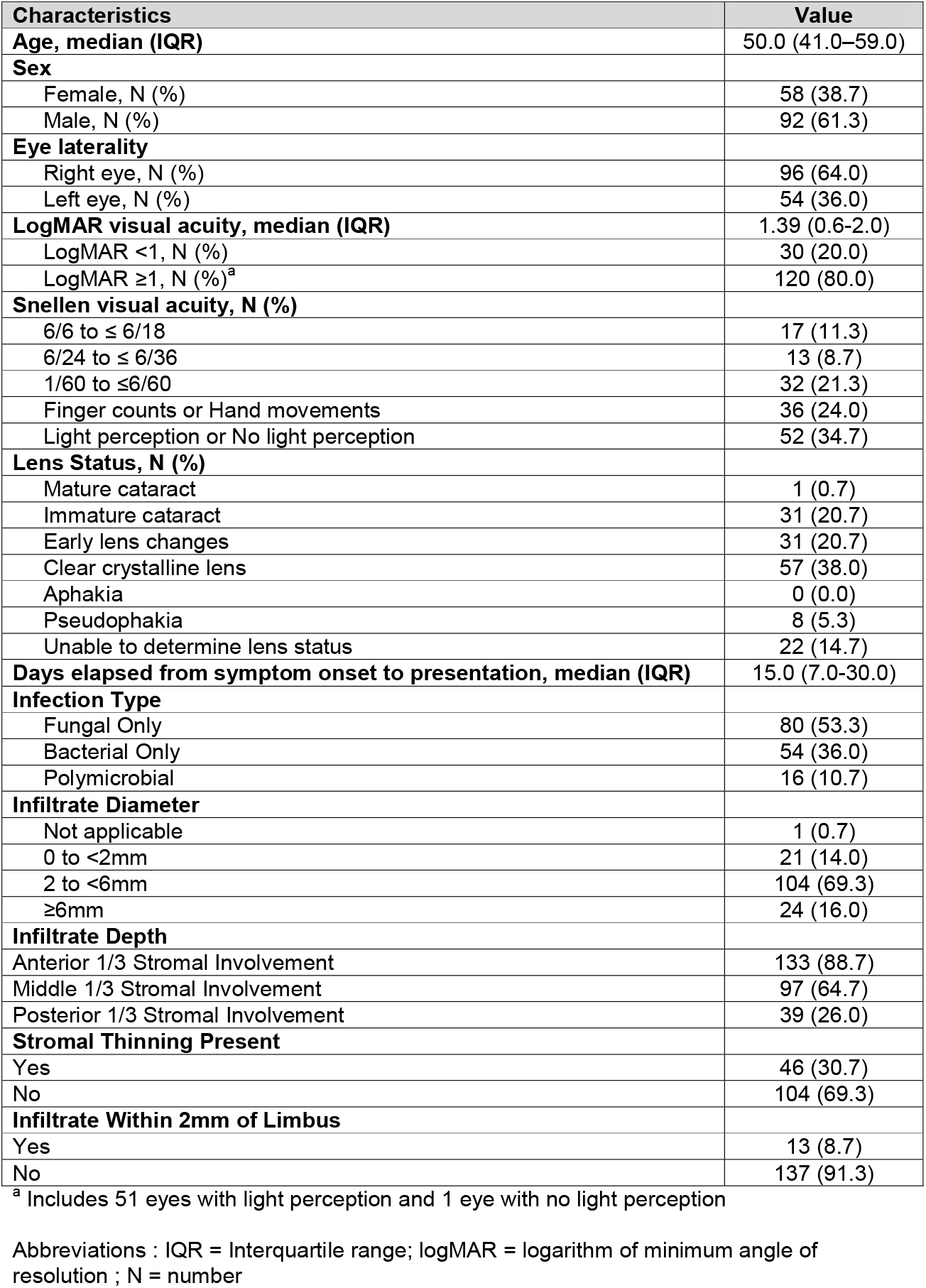
Demographic and clinical characteristics of study population.

| **Characteristics** | **Value** |
| --- | --- |
| **Age, median (IQR)** | 50.0 (41.0–59.0) |
| **Sex** |  |
| Female, N (%) | 58 (38.7) |
| Male, N (%) | 92 (61.3) |
| **Eye laterality** |  |
| Right eye, N (%) | 96 (64.0) |
| Left eye, N (%) | 54 (36.0) |
| **LogMAR visual acuity, median (IQR)** | 1.39 (0.6-2.0) |
| LogMAR <1, N (%) | 30 (20.0) |
| LogMAR ≥1, N (%)<sup>a</sup> | 120 (80.0) |
| **Snellen visual acuity, N (%)** |  |
| 6/6 to ≤ 6/18 | 17 (11.3) |
| 6/24 to ≤ 6/36 | 13 (8.7) |
| 1/60 to ≤6/60 | 32 (21.3) |
| Finger counts or Hand movements | 36 (24.0) |
| Light perception or No light perception | 52 (34.7) |
| **Lens Status, N (%)** |  |
| Mature cataract | 1 (0.7) |
| Immature cataract | 31 (20.7) |
| Early lens changes | 31 (20.7) |
| Clear crystalline lens | 57 (38.0) |
| Aphakia | 0 (0.0) |
| Pseudophakia | 8 (5.3) |
| Unable to determine lens status | 22 (14.7) |
| **Days elapsed from symptom onset to presentation, median (IQR)** | 15.0 (7.0-30.0) |
| **Infection Type** |  |
| Fungal Only | 80 (53.3) |
| Bacterial Only | 54 (36.0) |
| Polymicrobial | 16 (10.7) |
| **Infiltrate Diameter** |  |
| Not applicable | 1 (0.7) |
| 0 to <2mm | 21 (14.0) |
| 2 to <6mm | 104 (69.3) |
| ≥6mm | 24 (16.0) |
| **Infiltrate Depth** |  |
| Anterior 1/3 Stromal Involvement | 133 (88.7) |
| Middle 1/3 Stromal Involvement | 97 (64.7) |
| Posterior 1/3 Stromal Involvement | 39 (26.0) |
| **Stromal Thinning Present** |  |
| Yes | 46 (30.7) |
| No | 104 (69.3) |
| **Infiltrate Within 2mm of Limbus** |  |
| Yes | 13 (8.7) |
| No | 137 (91.3) |

<sup>a</sup> Includes 51 eyes with light perception and 1 eye with no light perception
Abbreviations : IQR = Interquartile range; logMAR = logarithm of minimum angle of resolution ; N = number

### Intra-Grader and Inter-Grader Agreement for ASOCT

Intra-grader agreement for perforation detection on ASOCT was excellent for both graders (Table 2). Among the 45 images evaluated in the repeat grading subsample, Grader 1 (NS) demonstrated 97.8% agreement (95% CI, 93.3-100.0%) with a kappa statistic of 0.92 (95% CI, 0.69-1.00). Grader 2 (FI) achieved 93.3% agreement (95% CI, 86.7-100.0%) with a kappa statistic of 0.76 (95% CI, 0.40-1.00). Examination of the grading matrices revealed that discordant assessments were relatively balanced: for Grader 1, 1 of 8 images (12.5%) initially graded as perforation present were subsequently graded as absent, while 0 of 37 images initially graded as absent were changed to present. For Grader 2, 2 of 8 images (25.0%) initially graded as perforation present were subsequently graded as absent, and 1 of 37 images (2.7%) initially graded as absent was changed to present (Table 2). Inter-grader agreement for perforation detection on ASOCT was near-perfect. Among 150 independently graded images, the two graders achieved 99.3% agreement (95% CI, 98.0-100.0%) with a kappa statistic of 0.98 (95% CI, 0.92-1.00). Only 1 of 150 images (0.7%) required adjudication due to disagreement between graders. Grader 1 identified perforation in 25 of 150 eyes (16.7%), while Grader 2 identified perforation in 24 of 150 eyes (16.0%). Following consensus adjudication, 24 eyes (16.0%) were classified as having frank perforation and 126 eyes (84.0%) were classified as non-perforated (Table 2).

**Table 2.** Intra-grader and Inter-grader Agreement of ASOCT and Slit Lamp Photography for Detection of Perforation.

|  |  | **ASOCT** | **Slit lamp**<br>**photography** |
| --- | --- | --- | --- |
| **Metric** | **N** | **Value (95% CI)** | **Value (95% CI)** |
| **Intra-grader agreement: Grader 1** | 45 |  |  |
| Percent Agreement |  | 97.8 (93.3-100.0) | 95.6 (88.9-100.0) |
| Kappa Statistic |  | 0.92 (0.69-1.00) | 0.85 (0.56-1.00) |
| **Intra-grader agreement: Grader 2** | 45 |  |  |
| Percent Agreement |  | 93.3 (86.7-100.0) | 91.1 (82.2-97.8) |
| Kappa Statistic |  | 0.76 (0.40-1.00) | 0.70 (0.36-0.94) |
| **Inter-grader agreement: Grader 1 vs 2** | 150 |  |  |
| Grader 1: Perforation Present, n (%) |  | 25 (16.7) | 18 (12.0) |
| Grader 2: Perforation Present, n (%) |  | 24 (16.0) | 19 (12.7) |
| Overall Percent Agreement |  | 99.3 (98.0-100.0) | 94.0 (90.0-97.3) |
| Kappa Statistic |  | 0.98 (0.92-1.00) | 0.72 (0.52-0.88) |

Abbreviations: ASOCT = anterior segment optical coherence tomography; CI = confidence interval; N = number

### Intra-Grader and Inter-Grader Agreement for Slit Lamp Photography

When evaluating eyes for presence or absence of perforation using slit lamp photographs alone, intra-grader agreement was substantial for both graders. Grader 1 had κ=0.85 (95% CI 0.56–1.00) with 95.6% agreement (95% CI 88.9–100.0), and Grader 2 had κ=0.70 (95% CI 0.36–0.94) with 91.1% agreement (95% CI 82.2–97.8) (Table 2). Inter-grader agreement was 94.0% (95% CI 90.0–97.3) with κ=0.72 (95% CI 0.52–0.88).

### ASOCT Versus Slit Lamp Examination

Consensus ASOCT grading identified perforation in 24 of 150 eyes (16.0%), whereas in-person slit lamp examination identified perforation in only 12 eyes (8.0%), representing a two-fold difference in detection using ASOCT. Overall agreement between ASOCT and in-person slit lamp examination was 86.6% (95% CI, 80.7-91.3%), with a kappa statistic of 0.38 (95% CI, 0.16-0.58) indicating fair agreement (Table 2).

Using ASOCT consensus grading as the reference standard, slit lamp examination demonstrated a sensitivity of only 33.3% (95% CI, 14.9-52.2%) for perforation detection, indicating that two-thirds of perforations identified on ASOCT were missed on slit lamp examination. Specificity was high at 96.8% (95% CI, 93.4-99.2%), indicating that slit lamp examination rarely identified perforation when ASOCT did not. The positive predictive value was 66.7% (95% CI, 37.5-92.0%) and negative predictive value was 88.4% (95% CI, 83.0-93.4%).

When the analysis was reversed to treat slit lamp examination as the reference standard, ASOCT demonstrated 100% sensitivity for detecting clinically apparent perforations, meaning that every perforation identified on slit lamp examination was also detected on ASOCT. However, specificity was correspondingly lower, reflecting the additional perforations detected by ASOCT that were not appreciated on slit lamp examination.

### Slit Lamp Photography Versus Slit Lamp Examination

Slit lamp photograph grading identified perforation in 19 of 150 images (12.7%). Using the in-person slit lamp examination as the reference standard, slit lamp photograph assessment yielded a sensitivity of 50.0% and a specificity of 90.6%. Agreement with the in-person diagnosis was fair (κ=0.32, 95% CI 0.09–0.54; percent agreement 87.3%, 95% CI 82.0–92.0) (Table 3).

**Table 3.**
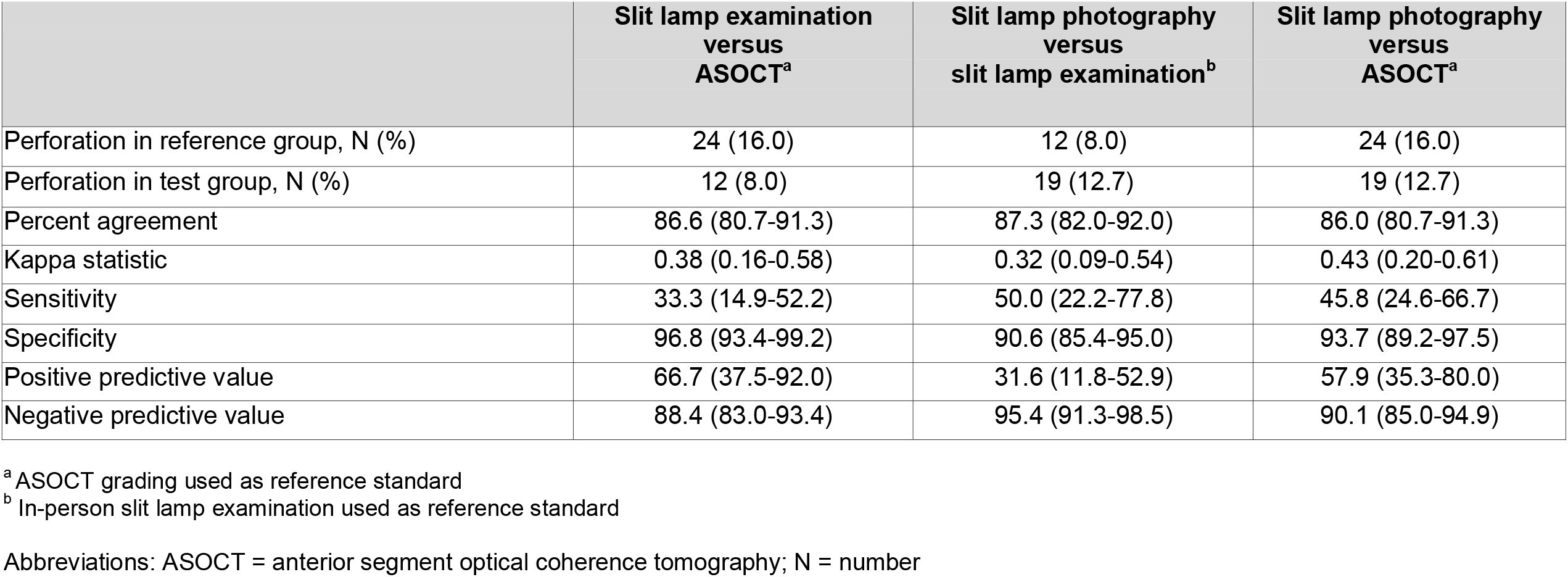
Concordance of ASOCT, Slit Lamp Photography, and In-Person Slit Lamp Examination for Detection of Perforation.

### ASOCT Versus Slit Lamp Photography

Using ASOCT composite image grading as the reference standard, slit lamp photography grading showed moderate concordance after accounting for chance agreement (κ=0.43) and an overall agreement of 86.0% (Table 3). The sensitivity was 45.8% and specificity 93.7%, with a positive predictive value of 57.9% and a negative predictive value of 90.1%.

### Factors Associated with Diagnostic Disagreement on ASOCT Versus Slit Lamp Examination

Logistic regression analysis identified several clinical factors associated with disagreement between ASOCT and slit lamp examination for detection of perforation (Table 4). Since ASOCT detected all cases of perforation identified on slit lamp examination plus additional cases, any disagreement between modalities reflects ASOCT detecting perforations missed clinically. In univariable analysis, disagreement was significantly higher for eyes with worse visual acuity (logMAR ≥1.0 vs. <1.0: OR=5.46; 95% CI, 0.70-42.8), indicating that ASOCT more frequently detected perforations missed by slit lamp examination in eyes with poorer vision. Eyes with infiltrates located in the middle one-third of the stroma had significantly higher odds of disagreement compared to eyes without mid-stromal involvement (OR=5.81; 95% CI, 1.29-26.24), and infiltrate location in the posterior one-third of the stroma was also associated with higher odds of disagreement (OR=4.45; 95% CI, 1.67-11.84). The presence of clinically evident stromal thinning on slit lamp examination was strongly associated with disagreement (OR=11.12; 95% CI, 3.24-38.27), as was bacterial compared to fungal infection (OR=6.02; 95% CI, 1.84-19.75). Infiltrates extending within 2mm of the limbus were also associated with higher odds of disagreement (OR=11.21; 95% CI, 3.24-38.27). In multivariable analysis adjusting for all covariates examined, mid-stromal involvement (OR=4.44; 95% CI, 1.08-18.30), stromal thinning (OR=8.19; 95% CI, 2.27-29.54), and infiltrate location within 2mm of limbus (OR=8.81; 95% CI, 1.77-43.80) remained independently associated with disagreement between ASOCT and in-person slit lamp examination (Table 4).

**Table 4.** Odds of Disagreement in Perforation Detection Between ASOCT and Slit Lamp Examination.

| Variable | Agreement, N (%) | Disagreement, N (%) | Unadjusted OR (95% CI)<sup>a</sup> | Adjusted OR (95% CI)<sup>b</sup> |
| --- | --- | --- | --- | --- |
| **Visual acuity** |  |  |  |  |
| LogMAR <1.0 | 29 (22.3) | 1 (5.0) | Reference | Reference |
| LogMAR ≥1.0 | 101 (77.7) | 19 (95.0) | 5.46 (0.70-42.8) | 7.70 (0.98-60.47) |
| **Infection type** |  |  |  |  |
| Fungal Only | 76 (58.5) | 4 (20.0) | Reference | Reference |
| Bacterial Only | 41 (31.5) | 13 (65.0) | 6.02 (1.84-19.75)<sup>c</sup> | 3.97 (0.93-16.87) |
| Polymicrobial | 13 (10.0) | 3 (15.0) | 4.38 (0.87-22.01) | 6.25 (0.99-39.30) |
| **Infiltrate diameter** |  |  |  |  |
| Not applicable | 1 (0.8) | 0 (0) | - | - |
| 0 to <2mm | 21 (16.2) | 2 (10.0) | Reference | Reference |
| 2 to <6mm | 89 (68.5) | 14 (70.0) | 1.65 (0.35-7.87) | 0.52 (0.08-3.20) |
| ≥6mm | 19 (14.7) | 4 (20.0) | 2.21 (0.36-13.55) | 0.41 (0.04-3.99) |
| **Infiltrate depth** |  |  |  |  |
| Anterior 1/3 stromal involvement | 116 (89.2) | 17 (85.0) | 0.68 (0.18-2.64) | 1.09 (0.15-7.91) |
| Middle 1/3 stromal involvement | 79 (60.8) | 18 (90.0) | 5.81 (1.29-26.24)<sup>c</sup> | 4.44 (1.08-18.30)<sup>c</sup> |
| Posterior 1/3 stromal involvement | 28 (21.5) | 11 (55.0) | 4.45 (1.67-11.84)<sup>c</sup> | 1.34 (0.38-4.64) |
| **Stromal thinning present on exam** |  |  |  |  |
| No | 99 (76.2) | 5 (25.0) | Reference | Reference |
| Yes | 31 (23.9) | 15 (75.0) | 9.58 (3.21-28.59)<sup>c</sup> | 8.19 (2.27-29.54)<sup>c</sup> |
| **Infiltrate within 2mm of limbus** |  |  |  |  |
| No | 124 (95.4) | 13 (65.0) | Reference | Reference |
| Yes | 6 (4.6) | 7 (35.0) | 11.12 (3.24-38.27)<sup>c</sup> | 8.81 (1.77-43.80)<sup>c</sup> |

<sup>a</sup> Unadjusted odds ratio for disagreement between ASOCT and slit lamp examination derived using univariable logistic regression. Because ASOCT detected all cases of perforation noted on slit lamp examination as well as additional cases, disagreement also indicates increased detection of perforation on ASOCT compared to slit lamp examination.
<sup>b</sup> Adjusted odds ratio derived using multivariable logistic regression after adjusting for visual acuity, infection type, infiltrate diameter, infiltrate depth, stromal thinning, and infiltrate proximity to limbus.
<sup>c</sup> P < 0.05. Displaying column percentages. Reference group for infiltrate depth was absence of the stromal involvement in respective categories
Abbreviations: ASOCT = anterior segment optical coherence tomography; CI = confidence interval; logMAR = logarithm of minimum angle of resolution; N = number; OR = odds ratio

## DISCUSSION

This study demonstrates that ASOCT enables highly objective and reproducible grading of corneal perforation in microbial keratitis and detects perforations with substantially greater sensitivity than in-person slit lamp examination. The diagnostic advantages of ASOCT were most pronounced in eyes with severe disease characterized by poor visual acuity, deep stromal involvement, stromal thinning, and bacterial etiology, which are precisely the clinical scenarios in which accurate perforation detection is most critical for guiding management decisions regarding observation, gluing, patching, or tectonic keratoplasty. Our findings have important implications for both clinical practice and the design of clinical research studies in microbial keratitis.

The excellent intra-grader and inter-grader concordance observed in this study provides strong evidence for the reliability of ASOCT-based perforation assessment. Intra-grader kappa statistics of 0.92 and 0.76 for the two graders, and an inter-grader kappa of 0.98, indicate excellent to near-perfect reproducibility^26^ despite the anatomic complexity and phenotypic variability of perforation presentations in microbial keratitis (Figure 1). Only 1 of 150 images required adjudication, underscoring the objectivity of ASOCT interpretation even when graders evaluated images independently without knowledge of each other’s assessments or clinical findings. This level of agreement suggests that ASOCT-based grading could provide a standardized, objective, and reproducible approach to perforation assessment that is not achievable with subjective slit lamp examination.

The finding that ASOCT detected twice as many perforations as in-person slit lamp examination (16.0% vs. 8.0%) and 25% more perforations than remote evaluation of slit lamp photographs (16.0% vs 12.7%) has significant clinical implications. Although surgical or histopathologic confirmation was not available, the additional perforations detected by ASOCT demonstrated anatomically plausible features including full-thickness stromal discontinuity, iris-cornea apposition at the site of thinnest cornea, and corresponding anterior chamber shallowing, which are findings that would be difficult to attribute to imaging artifact or overcalling of severe thinning alone. Pairwise agreement for perforation detection was greatest between ASOCT and slit lamp photography (κ=0.43), intermediate between ASOCT and in-person slit lamp examination (κ=0.38), and lowest between slit lamp photography and in-person slit lamp examination (κ=0.32). The low sensitivity of slit lamp examination (33.3%) for detecting perforations identified on ASOCT indicates that a majority of ASOCT-positive perforations may be clinically occult. Clinical detection of perforation at the slit lamp often relies on secondary signs such as Seidel positivity or frank anterior chamber collapse, whereas ASOCT enables direct visualization of full-thickness stromal disruption even when the perforation is small or plugged by iris tissue. The low sensitivity of slit lamp examination may therefore reflect not only the difficulty of examining opaque corneas but also a narrower clinical definition that requires more overt manifestations of perforation. In eyes with dense corneal opacification, stromal edema, or small perforation size, the biomicroscopic signs of perforation may be subtle or absent altogether, and even Seidel testing can be falsely negative when there is sufficient iris plugging. By providing high-resolution cross-sectional imaging of the corneal layers, ASOCT can visualize full-thickness discontinuity, anterior chamber shallowing, and iris-cornea contact that may not be apparent on external examination of an opaque cornea. This enhanced sensitivity is particularly valuable given the clinical importance of perforation as a risk factor for endophthalmitis and indication for urgent procedural intervention.

Our regression analysis revealed that ASOCT appears to provide the greatest incremental diagnostic value over slit lamp examination in precisely those eyes with the most severe disease where clinical detection of perforation is most challenging, such as eyes with poor visual acuity, deep stromal involvement, and advanced stromal thinning. This pattern likely reflects the greater difficulty of clinical examination in eyes with opaque, edematous corneas where direct visualization of the anterior chamber is impaired. For these challenging cases, ASOCT can penetrate opaque corneal tissue to reveal underlying structural abnormalities that escape detection on biomicroscopy. The association of bacterial infection with greater disagreement may relate to the more rapid progression and severe inflammation characteristic of bacterial compared to fungal keratitis, although this finding warrants further investigation and replication.

Slit lamp photography interpretation performed in a standardized manner by a panel of expert graders also enabled increased detection of perforation compared to slit lamp examination performed by several different ophthalmologists in a busy real-world clinical setting, highlighting the importance of using standardized imaging and image grading protocols in outcomes assessment for microbial keratitis. However, slit lamp photography was inferior to ASOCT for detection of perforation, likely because photographs lack OCT’s three-dimensional perspective and easier visualization of intraocular contents such as iris plugging or anterior chamber depth. This limitation has implications for clinical trial reading centers and telemedicine monitoring programs that rely on anterior segment photography. By providing standardized cross-sectional imaging, ASOCT may offer advantages over photography for remote perforation assessment.

### Clinical and Research Implications

Our findings have direct applications for clinical practice. In patients with severe microbial keratitis, particularly those with poor visual acuity and clinical features suggesting advanced disease, ASOCT imaging can serve as a useful adjunctive tool to detect perforations that may be missed on slit lamp examination. Early detection of perforation may prompt timely surgical intervention such as gluing, patching, or keratoplasty, more aggressive medical management, or closer monitoring to prevent progression to endophthalmitis. Conversely, the presence of occult or impending perforation could influence clinicians’ decision to avoid procedures such as intrastromal injections, intracameral injections, or anterior chamber washout which could enlarge pre-existing perforations. Early identification of high-risk cases using ASOCT could also allow surgeons to be better prepared for surgery, such as by obtaining therapeutic corneal tissue or tissue adhesive ahead of time for eyes at high risk of developing large perforation intraoperatively. Such considerations are particularly relevant during evaluation of infections with dense infiltrates, edema, or endothelial plaque which can preclude visualization of perforation.

The non-contact nature of ASOCT and its ability to image through corneal opacity make it particularly well-suited for assessing fragile or severely compromised corneas where examination under bright slit lamp illumination may be uncomfortable or technically difficult. Additionally, the telemedicine potential of ASOCT imaging could enable remote expert consultation for perforation assessment in settings without immediate access to cornea specialists, potentially improving care delivery in resource-limited environments where microbial keratitis burden is highest.

This study’s findings also carry implications for the design and conduct of clinical trials in microbial keratitis. Perforation is frequently employed as a key efficacy endpoint or adverse event in trials evaluating antimicrobial therapies, adjunctive treatments, and surgical interventions, as demonstrated by the Mycotic Ulcer Treatment Trial.^14,15^ The subjective nature of slit lamp-based perforation assessment introduces measurement variability that may be compounded in multicenter trials with multiple examiners at different sites. Such variability reduces statistical precision and may obscure true treatment effects. ASOCT-based grading, with its demonstrated high inter-grader concordance, offers a pathway to standardize perforation assessment across trial sites. Implementation of a centralized image reading center, where masked graders evaluate ASOCT images using standardized criteria, would provide objective, reproducible outcome ascertainment regardless of which clinician treated the patient at each individual site. This approach reduces ascertainment variability, increases statistical power, and ultimately improves the ability to detect meaningful differences between treatment groups, facilitating the rigorous evidence generation needed to advance care for microbial keratitis.

### Strengths and Limitations

This study has several strengths. The prospective design with consecutive patient enrollment minimizes selection bias, and the sample size of 150 eyes exceeds that of prior studies examining ASOCT for corneal perforation assessment in microbial keratitis. The rigorous methodology, including masked independent grading by two experienced ophthalmologists, repeat grading of a substantial subsample to assess reproducibility, and consensus adjudication of disagreements, provides robust estimates of diagnostic concordance. The inclusion of detailed clinical characterization allowed identification of specific factors associated with discordance between imaging and clinical assessment.

Several limitations merit consideration. First, slit lamp examination was performed by fully trained ophthalmologists but was not performed by the same specialists in all cases. Use of Seidel testing during slit lamp examination was not required nor recorded. Future research should evaluate multiple masked clinicians’ diagnostic agreement in gauging features such as perforation or percentage stromal thinning in the same eye. As clinicians show considerable variability in measurement of easier-to-visualize features such as epithelial defect size,^27,28^ some variability in measurement of stromal thinning or perforation may also be expected, and this may have led to inconsistency in diagnostic performance of the “gold standard” of slit lamp examination. The logistical infeasibility of having multiple ophthalmologists perform in-person examination for each patient precluded assessment of inter-examiner variability for slit lamp-based perforation detection. Based on clinical experience and the recognized subjectivity of perforation assessment, we expect that inter-examiner variability for slit lamp examination would be considerably higher than the near-perfect inter-grader agreement observed for ASOCT.

Second, although all examining ophthalmologists were required to note the presence of frank perforation, we did not systematically record data on iris plugging or anterior chamber depth, which could have allowed further comparison between in-person examination and imaging. This study also focused on detection of frank perforation, a typically obvious finding on ASOCT imaging. We did not systematically evaluate ASOCT for detection of impending perforations, descemetoceles, healed old perforations, or other anterior chamber abnormalities such as iridocorneal adhesions that could also guide clinical or surgical decision-making., ASOCT’s diagnostic superiority over slit lamp examination for detection of frank perforations may mean that ASOCT is also superior for detecting subtler forms of perforation. Future studies should evaluate ASOCT’s ability to detect such abnormalities using agreed-upon criteria.

Third, this study did not directly compare detection of perforation via slit lamp photography versus ASOCT alone, but rather compared in-person slit lamp examination to composite images that included both slit lamp photographs and ASOCT scans. Slit lamp photographs only included diffuse illumination and blue light images and did not include slit beam images or sclerotic scatter, which might have improved detection of stromal thinning, perforation, and anterior chamber collapse. Nevertheless, the excellent diagnostic performance of ASOCT grading and the relatively lower sensitivity of slit lamp examination suggest that ASOCT alone is likely superior to slit lamp photography for detecting perforation, particularly for remote assessment.

Fourth, the ASOCT machine used in this study (Heidelberg Anterion) offers limbus-to-limbus imaging and deeper tissue visualization compared with other devices such as the Heidelberg Spectralis and Zeiss Cirrus, and this study’s results may therefore not be directly generalizable to settings using different ASOCT platforms. This study evaluated images taken with the Anterion’s Metrics App, which obtains six radial cross-sections of the cornea. Results may differ with other Anterion settings such as the Imaging application, which enables higher sampling density via radial or raster scans, or raster scans that have shorter scan length and do not image the entire cornea in a limbus-to-limbus fashion. If another ASOCT machine with comparable scan length obtained greater than six cuts through the cornea, it could potentially have even more sensitive detection of perforation and iris abnormalities.

Fifth, as this study was conducted at a single high-volume eye hospital in India with experienced graders and standardized imaging protocols, these findings should be replicated in other populations, healthcare settings, and imaging devices. Finally, we also did not longitudinally track clinical outcomes among eyes with ASOCT-detected perforations that were missed on slit lamp examination. Future studies correlating these imaging findings with subsequent need for surgical intervention would help establish their clinical significance.

## CONCLUSION

In conclusion, ASOCT detected corneal perforation in microbial keratitis with greater sensitivity and comparable specificity compared to in-person slit lamp examination, enabling human grading of perforation with high concordance and reproducibility. ASOCT detects perforations with far greater sensitivity than clinical examination, particularly in eyes with more severe disease and opaque corneas. These findings support the use of ASOCT as an objective, standardized tool to improve clinical assessment of microbial keratitis and to enhance the quality and rigor of outcome ascertainment in future clinical trials.

## Data Availability

Data produced in the present study may be made available upon reasonable request to the authors and in accordance with local data sharing regulations.

## ACKNOWLEDGMENTS

The authors thank the clinical and research staff at SNC Chitrakoot for their assistance with patient recruitment and data collection.

## FUNDING

This work was supported by the National Institutes of Health (K23EY032988 and R33EY034343 to N.S.S.) and KeraLink International. The funding organizations had no role in the design or conduct of this research.

## DISCLOSURES

The authors declare no financial conflicts of interest related to this work.

## Notes

### Competing Interest Statement

The authors have declared no competing interest.

### Author Declarations

The IRB of the Johns Hopkins University School of Medicine gave ethical approval for this work. The ethics committee SNC Chitrakoot Eye Hospital gave ethical approval for this work.

